# Asymptomatic hypoglycemia among preterm newborns: a cross-sectional analysis

**DOI:** 10.1101/2023.12.30.23300665

**Authors:** Shani S. Salum, Florence S. Kalabamu, Maulidi R. Fataki, Salha A. Omary, Ummulkheir H. Mohammed, Hillary A. Kizwi, Kelvin M. Leshabari

## Abstract

**Background:** Hypoglycemia is the commonest metabolic abnormality encountered in newborns. Preterm newborns are the group with the highest risk of clinically significant hypoglycemia. However exact factors associated with asymptomatic neonatal hypoglycemia is not known.

**Objective:** To assess factors associated with asymptomatic hypoglycemia among preterm newborns.

**Materials and Methods:** A cross sectional hospital analytical-based study was carried out at Dar es salaam public regional referral hospitals. Specifically, the study was done at Amana, Mwananyamala and Temeke. Data on demographic and clinical characteristics of preterm newborns and their mothers were collected and analyzed using Epi-Info™ software version 7.4. Main data analysis was done by applying a multivariable logistic regression model with neonatal random glycaemia coded in a binary fashion at a cut-off point of 2.6 mmol/L. An α-level of 5% was used as a limit of type I error.

**Results:** We recruited and analysed 217 asymptomatic preterm newborns within 6-24 hours post-delivery. Male: Female = 1.1:1 (females n=105, 48.4%). Median glycemic level was 2.6 (IQR; 2.1-3.9) mmol/L. Breastfeeding within 1st hour post-delivery was the only statistically significant protective factor against development of neonatal hypoglycemia (OR; 0.123, 95%-CI; 0.052-0.287) in a fitted multivariable logistic regression model.

**Conclusion:** Nearly half of all asymptomatic newborns were in hypoglycemic range. Delayed breast feeding within 1^st^ hour post-delivery was associated with neonatal hypoglycemia.

**Recommendations:** Breastfeeding practices within 1^st^ hour post-delivery need to be emphasized to all expectant mothers in order to avoid risk of asymptomatic neonatal hypoglycemia. Preterm neonates maybe the risky group worth considering for continuous glycemic screening at hospital protocols.

## Introduction

Hypoglycemia is one of the most prevalent metabolic abnormalities encountered in preterm newborn babies (1). However, hypoglycemia in newborns is still a matter of controversy to date. Controversies are both quantitative as well as qualitative. For instance, the glycaemic cut-off point of less than 2.6 mmol/L (in US – 50mg/dL equivalent to 2.75mmol/L) still raises concerns to practicing neonatologists (2). Globally, those on quantitative reasoning do consider the magic number (2.6 mmol/L) as the cornerstone of measurements and standardization. Qualitatively, others question the practicality of the set magic number for diagnosing neonatal hypoglycaemia. Scholars antagonising the concept consider the argument on possible evolutionary basis for its occurrence within the first 24-hours of extra-uterine life. Scholars justified their thinking on the nosology to be based on symptoms and signs (clinical aspects), since for that particular condition, quite often clinicians cannot differentiate physiology from possible pathology using figures alone. Thus, on the basis of qualitative aspects, it is still unclear whether the agreed cut-off point for neonatal hypoglycaemia has a universal clinical utility in neonatology. (2)

Prolonged, neonatal hypoglycemia has been shown to potentiate mental retardation, neurological deficits and recurrent seizures (3) Some studies have shown perinatal hypoxia, small for gestational age and maternal hypertension to be factors associated with hypoglycemia in preterm newborns (4, 5) However, the exact mechanisms behind the observations are yet to be identified by neonatologists/endocrinologists to date. Yet still, it is not clear whether the suggested risks are generalized or specific to a certain neonatal cohort alone. At present, there is no evidence from available/retrievable published databases of any findings on factors associated with neonatal hypoglycemia among preterm babies in Tanzania. Besides, for findings published from elsewhere, almost none assessed the concept of hypoglycemia among asymptomatic neonates.

## Methods

A cross sectional hospital analytical-based study was carried out at Dar es Salaam public regional referral hospitals. Specifically, the study was done at Amana, Mwananyamala and Temeke regional referral hospitals. The three hospitals are the registered public regional referral facilities for Ilala, Kinondoni and Temeke municipalities in Dar es Salaam, Tanzania. We believed the hospitals to be receiving a representative sample of Dar es Salaam residents.

We planned and recruited all preterm newborns found at neonatal wards and Neonatal Intensive Care Unit (NICU) within 6-24 hours of postnatal life at the three hospitals during the study period. Specifically, the study commenced in June 2022, and run up to (and including) November 2022. A minimum sample of 198 preterm neonates were required in order to achieve a study power of 80% under the assumption of 5% α-level for disproving type 1 error rates in findings. Preterm newborns with asymptomatic hypoglycemia were the target population. Newborns either with congenital malformations, referred from lower facilities due to other neonatal complications, home delivered or babies symptomatic for hypoglycemia were excluded. Data was collected using a structured questionnaire, after receipt of a written informed consent, to participate in the study, from post-partum mothers. Mothers of preterm newborns were interviewed and their immediate past gestational age assessed using Ballard score chart. Nutritional status was assessed using growth charts. Random (venous) blood glucose of the preterm newborns screened using Glucoplus blood glucose monitors (Glucoplus™ Inc. 2004, Canada). Principal Investigator and trained research assistants (neonatal ward nurses) were involved in data collection using the tools (Questionnaire, Ballard score chart, growth chart, Glucoplus™) Both principal investigator and nurses underwent formal pilot practice for 1 day prior to actual use of the tool.

Immediately following data collection, data cleaning followed at the end of each day. The exercise was done by the principal investigator herself. It mainly involved checking for consistency in responses given, screening for recording errors as well as coding-decoding of questionnaire responses. Data was triple entered under the supervision of the Principal Investigator and data scientist. Data was then stored by principal investigator until analysis time. Data was analysed using Epi-Info™ statistical software version 7.4 (CDC, Atlanta - USA). Descriptive statistics were summarized as median with corresponding inter-quartile range (for quantitative variables) or frequency and proportion (for qualitative variables). Initial data analysis involved exploratory data analysis and mainly screened data for possible linearity/homoscedasticity/normality/non-autocorrelation assumptions between dependent (asymptomatic hypoglycemia) and independent variables (series of clinical and demographic risk factors), outlier analysis, box plots and scatter plots as well as logistic function between outcome variable (hypoglycemia – coded as 1-if random glucose value was < 2.6 mmol/L and 0 – if random glycaemia was ≥ 2.6 mmol/L). Factors associated with hypoglycemia were assessed using binary multivariable logistic regression model. An alpha level of 5% was used as a limit against type I error in findings.

Ethical approval was sought from Hubert Kairuki Memorial University’s Institutional Research Ethical Committee as well as the Regional Referral Hospital Authority, which included the Assistant Executive Director of Amana Regional Referral Hospital, the Medical Officer in Charge of Mwananyamala Regional Referral Hospital, and the Medical Officer in Charge of Temeke Regional Referral Hospital. Written informed consent to mothers included description about the study goal, risks and benefits of inclusion of their newborns into the study as well as awareness raising on the voluntary nature of participation into the study. Besides, a note that participation/withdrawal from the study had minimal influence on the planned care at the sites was also part of the written informed consent form. The study was designed to have minimal risks like pain, bleeding, swelling on prick site. These were controlled by applying pressure compression on the prick site. For those preterm newborns found with low blood glucose level, the study team in collaboration with Team providing care in the ward promptly initiated appropriate treatment by administering intravenous 10% dextrose 2mls/kg bolus followed by intravenous infusion and close follow-up to prevent rebound hypoglycemia as per local hospital protocols for management of neonatal hypoglycaemia.

## Results

We recruited and analysed 217 preterm neonates asymptomatic for hypoglycemia at Dar es Salaam Regional Referral Hospitals from June –Nov 2022. They constituted 100, 67 and 50 preterm newborns from Amana, Temeke and Mwananyamala Regional Referral Hospitals respectively. The baseline characteristics of study participants are as summarised in table 1 below:

**Table 1:**
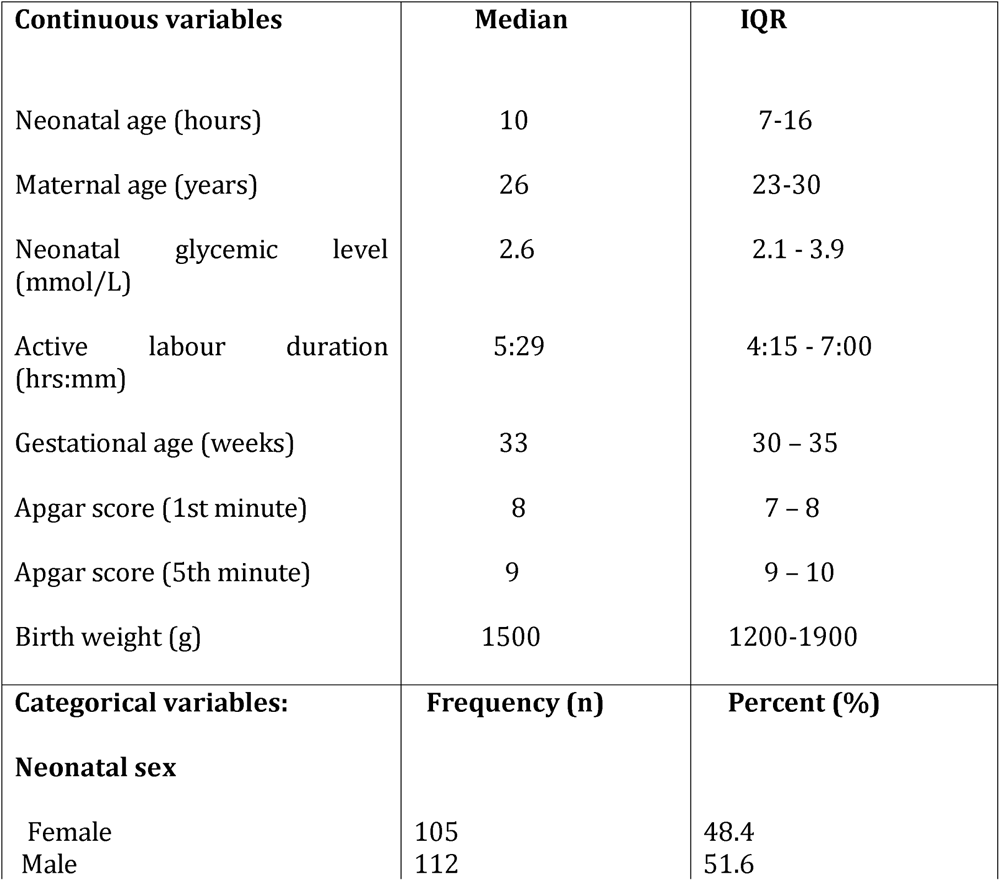
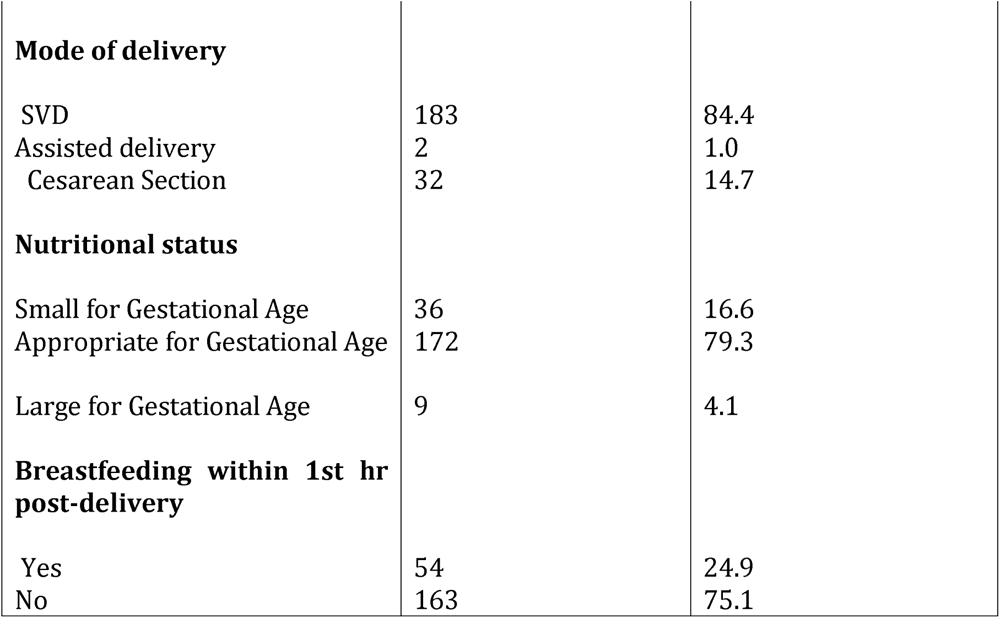
Baseline characteristics of selected maternal, neonatal and institutional factors associated with asymptomatic hypoglycemia among preterm newborns seen in Dar es salaam public regional referral hospitals.

**TABLE 2:**
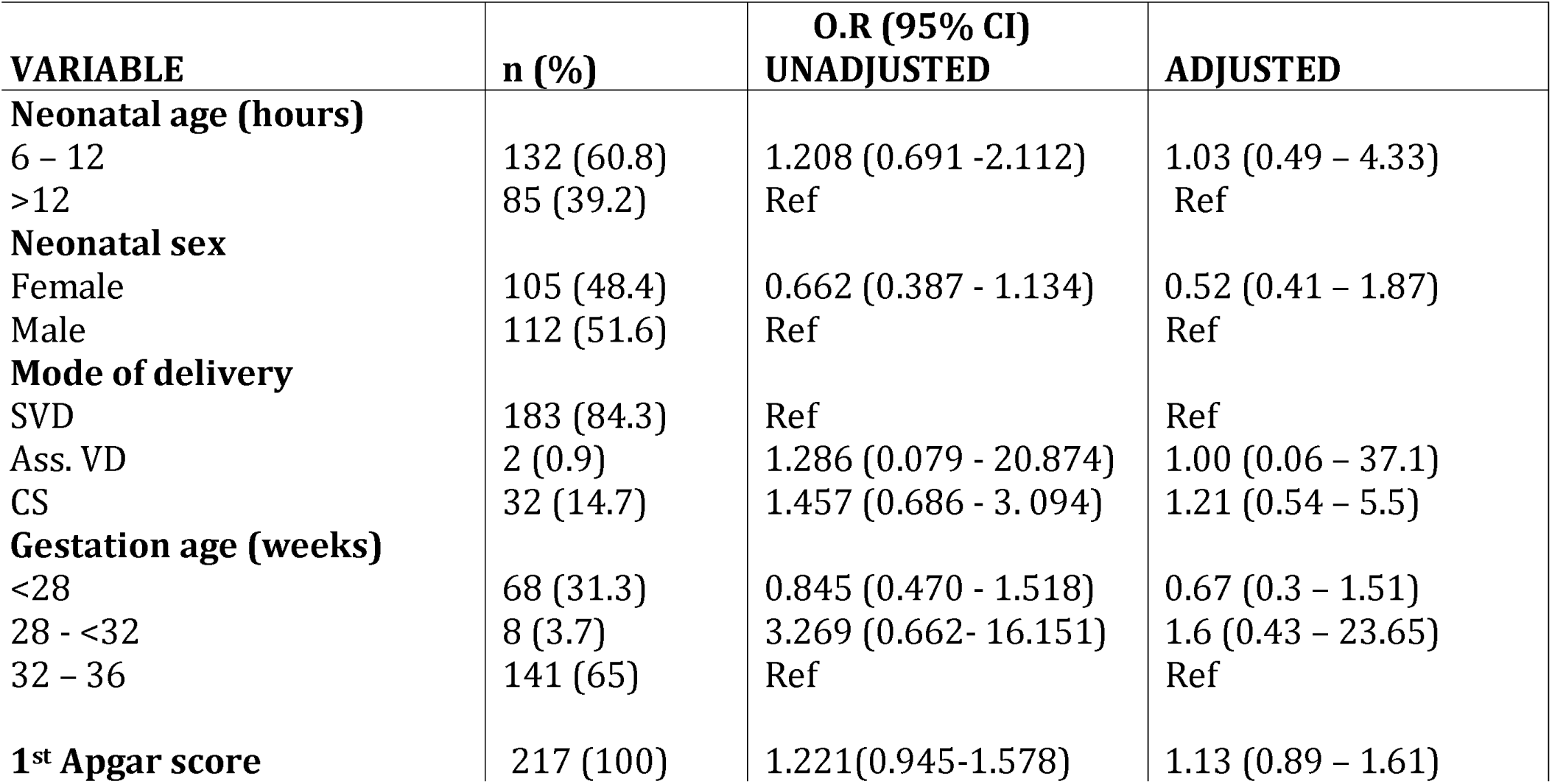
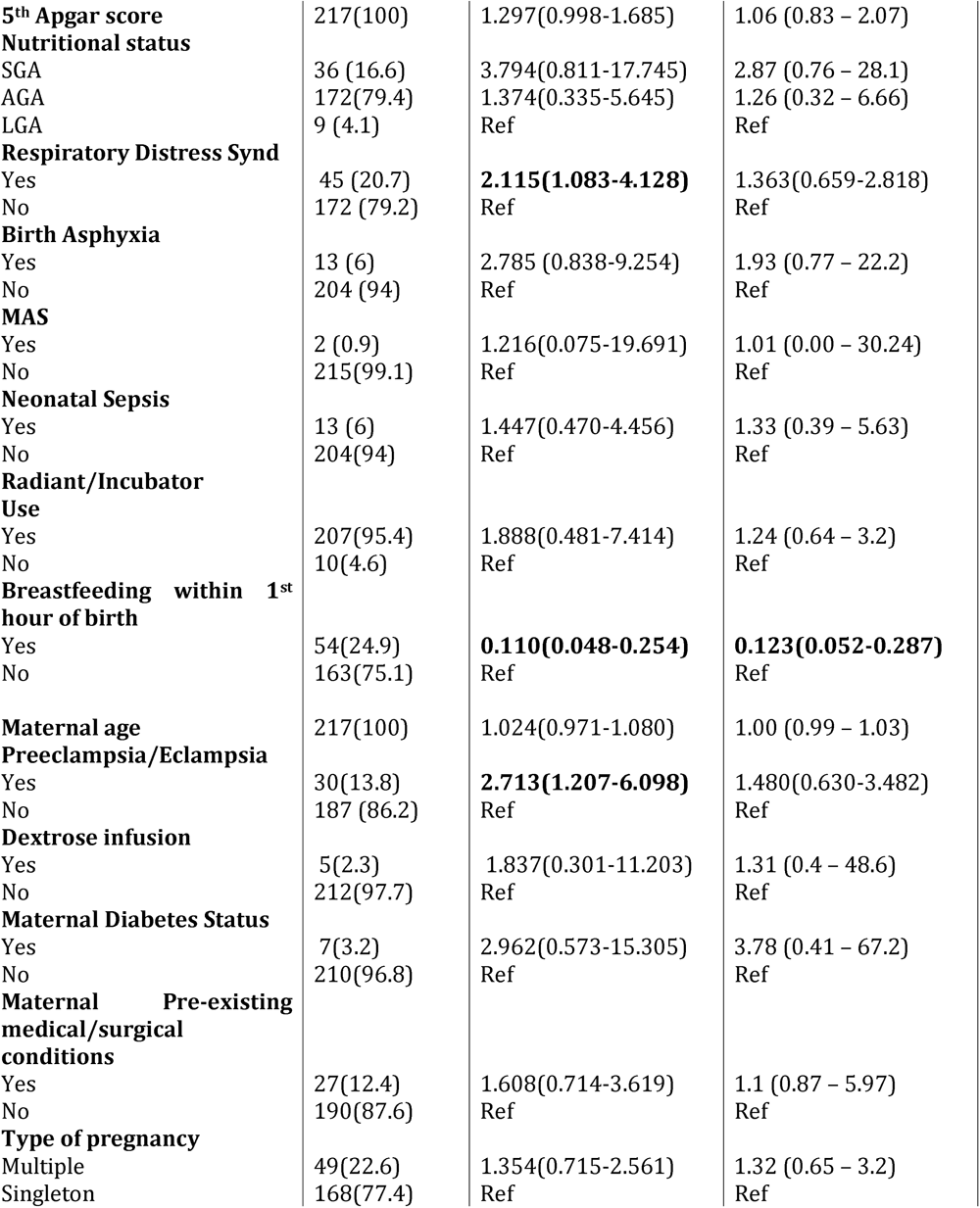
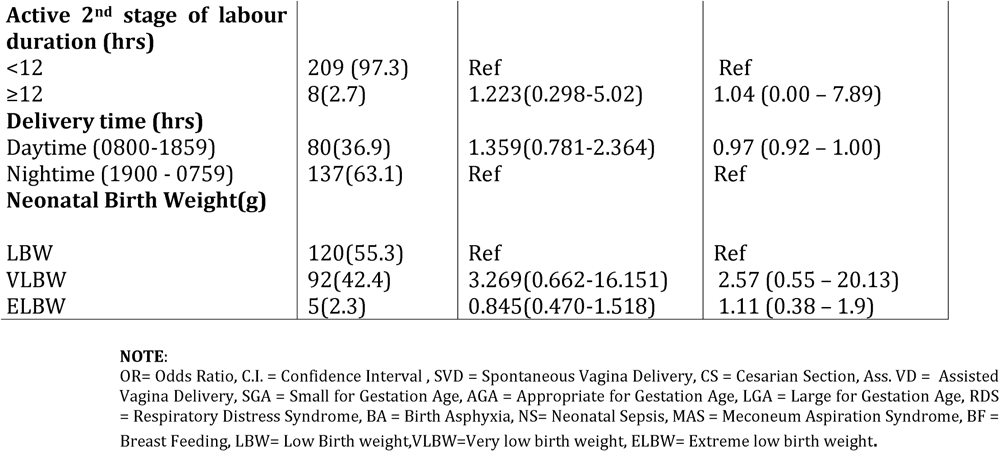
Univariate and multivariable binary logistic regression analysis of selected clinical factors associated with asymptomatic hypoglycemia among preterm newborns seen at Dar es salaam public regional referral hospitals.

## DISCUSSION

In this study, we found almost half of all preterm babies to have been in quantitative hypoglycemic range. It is a matter of special interest to realize that this estimate was derived out of asymptomatic preterm babies. In view of the fact that the figures happened to preterm delivered babies, the common understanding that neonatal hypoglycemia needs to be intervened clinically only upon symptoms need an additional thought to decision makers in health, and especially neonatologists in resource limited settings. Specifically, it appears preterm delivered babies are potential risky group among neonates for development of hypoglycaemia. Thus, hospital policies/protocols and national guidelines may consider these findings as potential signals towards including this specific neonatal group for urgent/frequent glycaemic screening. The current findings are comparable to others in similar settings done before (7–10) For instance, Sultan and his colleagues found out 73% of all babies born prematurely in South-Eastern Tanzania to had been in hypoglycemic range (7). However, the study is about two decades old and no newer study could be retrieved from peer reviewed platforms on the topic. It was a large population-based survey done among neonates in South-Eastern Tanzania back in 2003-2007(7) At a gross scale, in addition to both findings displaying prevalent hypoglycaemia in neonates, there are a number of factors behind the significant differences between our study and Sultan’s study. For instance, whereas we only considered asymptomatic preterm newborns, Sultan’s study included all neonates. Besides, Sultan’s study had a longer study duration, and therefore, better likelihood to capture a more robust estimate unlike ours. Otherwise, in the same East African zone, there have been some other notable differences in findings to ours. For instance, Mukunya and others in their Uganda’s community-based study found a prevalence of 2.2% for neonatal hypoglycemia in their screening exercise. (11) The fact that this metabolic abnormality seems prevalent in Africa, and especially Tanzania, is a wake-up call for urgent policy and guidelines changes on the construct of neonatal hypoglycemia. It is highly likely that the condition is a silent trigger to other neonatal ill-states and even a risk for other maladies later in adult life, hence prompt diagnosis and management is highly warranted.

Accordingly, the median random blood glucose of 2.6mmol/l among our study population are likely to be justifiable evidence for glycemic screening even among asymptomatic neonates. There are several mechanisms that accounts for possibilities of the metabolic abnormality in neonates. The factors of limited glycogen and fat stores, inability to generate new glucose using gluconeogenesis pathways, with higher metabolic demands due to a relatively larger brain size, and hyperinsulinemia are among the known etiological factors for the evolution of neonatal hypoglycemia. (11) However, our study was cross-sectional by design, and hence we could not make follow-up data of the glycemic ranges in studied children nor could we justify the single glycemic output to be significant to justify neonatal hypoglycemia in a clinical sense. However, given our study findings, we can safely deduce that glycemic checks need to be instituted to all neonates regardless of their initial clinical stata.

We also tested a number of known associated factors for neonatal hypoglycemia but found breastfeeding within 1st hour of birth to be the only clinically meaningful factor in preventing neonatal hypoglycemia in asymptomatic phase. Likewise, Mukunya and others in their Uganda’s community-based study found that delayed breastfeeding initiation was contributing neonatal hypoglycemia (11). Breastfeeding was found to be the most significant factor protective against hypoglycemia in our study. Breastfeeding has been evidently associated with glucose stability in the 1^st^ 24 hours of life on earth previously among term infants. (12, 13). Otherwise, our findings are consistent and reflective of what was seen before in a large quasi-experimental study from Denmark. (14) Accordingly, the Danish study assessed the contribution of early breastfeeding to prevention of neonatal hypoglycemia using skin-to-skin contact un-interruptive for at least 2-hours immediately post-delivery. (14) In contrast, our study was cross-sectional by design. Therefore, exact time data on breastfeeding initiation could not be assessed as it lied beyond our study objectives. Moreover, our study could not manage to assess temporal relationship between breastfeeding and incident hypoglycemia in asymptomatic neonates. Besides, there is no evidence of any statistically significant data for similar studies in East Africa and even sub-Saharan Africa. Thus, future studies on this topic needs to be designed as interventional and quantitative in nature in order to arrive with numerical estimates for comparison purposes.

Our study findings are unique as they report asymptomatic neonatal hypoglycemia and through analysis, we managed to confer potential solution to the clinical challenge. Moreover, our study findings included maternal, institutional as well as neonatal factors known to be associated with neonatal hypoglycemia in asymptomatic preterm babies. Thus, we believe our findings can provisionally be used by policy makers in decision making process while waiting for more prospective designed evidence in future. However, the study was limited in time (6-24 hours) of glycemic assessment in early neonatal period. The decision to limit the time is subject to earlier observations on the probable physiologic basis of glucose homeostasis early in extra-uterine life (15–17). Thus, should there be any deviations in glycemic state over time, our study was limited to capture such an information.

In a pioneering move, our study findings highly suggest possibilities of positive benefits of breastfeeding within 1^st^ hour post-delivery to preterm newborns. Care needs to be taken to expectant mothers using safe-motherhood initiatives. Exclusively breastfeeding within 1 hour of birth should be emphasised as they were significantly protective against neonatal hypoglycemic range. On a strict evolutionary sense, there appears to be potential utility of low blood sugar levels to newborns immediately upon entry to extra-uterine life (18). To what extent do these findings translate to prevalent double burden of morbidity and mortality indices among under-five year population in Tanzania and beyond, is still a matter of speculation (25, 26). However, the fact that there is solid evidence for under-fives population to be in ill-state in Tanzania (22) calls for urgent considerations for more analytical, possibly longitudinal designs in ascertaining the long-term effects of asymptomatic hypoglycemia in newborns. Besides, these findings call for urgent and reliable estimates of morbidity and mortality statistics of special groups in the population pyramid. At present, there are potential indications that shows certain segments of the Tanzania’s population are left out in most reported morbidity and mortality statistics (25, 26). There are potential clues that chronic ill-states in childhood may result to miserable later life years both in Africa and elsewhere (23, 24, 27). As we are advancing medical sciences on ageing cascades, even among humans (19, 24), we wish to consider that as a treasure hunt at least for now.

## Data Availability

All data produced in the present study are available upon reasonable request to the authors

https://www.medrxiv.org/content/10.1101/2022.10.28.22281650v1.full.pdf

